# A machine-learning model to harmonize brain volumetric data for quantitative neuro-radiological assessment of Alzheimer’s disease

**DOI:** 10.1101/2024.02.01.24302048

**Authors:** Damiano Archetti, Vikram Venkatraghavan, Béla Weiss, Pierrick Bourgeat, Tibor Auer, Zoltán Vidnyánszky, Stanley Durrleman, Wiesje M. van der Flier, Frederik Barkhof, Daniel C. Alexander, Andre Altmann, Alberto Redolfi, Betty M. Tijms, Neil P. Oxtoby, Australian Imaging Biomarkers and Lifestyle Study, Alzheimer’s Disease Neuroimaging Initiative, the E-DADS Consortium

**Author notes:** **Corresponding author:** Damiano Archetti, Address: Via Pilastroni 4, Brescia, 25125, Italy. Indicates shared first co-authorship, with the first co-authors being listed alphabetically. Data used in the preparation of this article was obtained from the Australian Imaging Biomarkers and Lifestyle (AIBL) Study. Unless named, the AIBL researchers contributed data but did not participate in analysis or writing of this report. AIBL researchers are listed at https://aibl.org.au. Data used in preparation of this article were obtained from the Alzheimer’s Disease Neuroimaging Initiative (ADNI) database (adni.loni.usc.edu). As such, the investigators within the ADNI contributed to the design and implementation of ADNI and/or provided data but did not participate in analysis or writing of this report. A complete listing of ADNI investigators can be found at: http://adni.loni.usc.edu/wp-content/uploads/how_to_apply/ADNI_Acknowledgement_List.pdf. **Contributors’ contacts** Vikram Venkatraghavan, Béla Weiss, Pierrick Bourgeat, Tibor Auer, Zoltán Vidnyánszky, Stanley Durrleman, Wiesje M. van der Flier, Frederik Barkhof, Daniel C Alexander, Andre Altmann, Alberto Redolfi, Betty M. Tijms, Neil P Oxtoby.

## Abstract

**Background:** Structural MRI plays a pivotal role in the radiological workup for assessing neurodegeneration. Scanner-related differences hinder quantitative neuroradiological assessment of Alzheimer’s disease (QNAD). This study aims to train a machine-learning model to harmonize brain volumetric data of patients not encountered during model training.

**Method:** Neuroharmony is a recently developed method that uses image quality metrics (IQM) as predictors to remove scanner-related effects in brain-volumetric data using random forest regression. To account for the interactions between AD-pathology and IQM during harmonization, we developed a multi-class extension of Neuroharmony. We performed cross-validation experiments to benchmark performance against existing approaches using data from 20,864 participants comprising cognitively unimpaired (CU) and impaired (CI) individuals, spanning 11 cohorts and 43 scanners. Evaluation metrics assessed ability to remove scanner-related variations in brain volumes (biomarker concordance), while retaining the ability to delineate different diagnostic groups (preserving disease-related signal).

**Results:** For each strategy, biomarker concordances between scanners were significantly better (p < 10^−6^) compared to pre-harmonized data. The proposed multi-class model achieved significantly higher concordance than the Neuroharmony model trained on CU individuals (CI: p < 10^−6^, CU: p = 0.02) and preserved disease-related signal better than the Neuroharmony model trained on all individuals without our proposed extension (ΔAUC= −0.09). The biomarker concordance was better in scanners seen during training (concordance > 97%) than unseen (concordance < 79%), independent of cognitive status.

**Conclusion:** In a large-scale multi-center dataset, our proposed multi-class Neuroharmony model outperformed other strategies available for harmonizing brain-volumetric data in a clinical setting. This paves the way for enabling QNAD in the future.

## 1 Introduction

Structural MRI such as T1-weighted (T1w) sequences are routinely acquired in memory clinics for diagnosing Alzheimer’s disease (AD)^1^, clinical phenotyping^2^, and for differentiating AD from other types of dementias^3^. In current clinical practice, radiologists primarily assess global and regional atrophy through visual examination of MRI. However, visual examinations are subjective and prone to intra-rater and inter-rater variability. Quantitative imaging biomarkers such as brain volumetric data are becoming increasingly popular because of their potential to improve diagnostic confidence^4^. Quantitative imaging biomarkers can be used for objective assessment in the radiological workflow either by using automated digital tools based on normative modelling^3^ or using latest advances in artificial intelligence such as brain-age estimation^5^, or data-driven subtyping^6^.

However, differences in MRI acquisition protocols and scanners affect consistency and reproducibility of brain volumetry^7^ and are a major impediment for the clinical translation to automated tools. To tackle this problem, many harmonization tools have emerged in recent years^8^. Such algorithms can either harmonize original scans^9^ or derivatives extracted from the scans^10^. Some of these algorithms have been shown to harmonize patient data affected by a neurodegenerative disease^11,12^, while preserving disease-related signature. However, such harmonization techniques typically work only for the scanner models they have been trained on, and, in some instances require the same subjects to be scanned with multiple different scanners^13^. Harmonizing volumetric data from MRI scanners not encountered during initial model training needs additional training with a substantial number of images from these scanners^14^. This poses a challenge for the deployment of such methods for clinical use.

Neuroharmony^15^ is a recently developed harmonization approach that can harmonize volumetric data from new and unseen MRI scanners. It works under the assumption that the corrections needed to harmonize data from multiple scanners can be predicted from image quality metrics (IQM) computed from the scans. While the original experiments indicate that harmonization works for healthy controls, harmonizing data from patients with neurodegenerative diseases remains an open problem. This is because disease pathology in patients may affect the IQM, and such effects remain unaccounted for in a Neuroharmony model trained on healthy controls.

In this paper we propose an extension of Neuroharmony to account for interaction between disease pathology and IQM to remove scanner-related effects (multi-class model of Neuroharmony). We systematically compare the performances of the proposed multi-class model in harmonizing data with two other approaches: the original Neuroharmony model trained only on cognitively unimpaired (CU) individuals (normative model of Neuroharmony) and the original Neuroharmony model trained on cognitively unimpaired as well as cognitively impaired (CI) individuals without our proposed multi-class extension (inclusive model of Neuroharmony). We used data from 11 cohorts across three continents for evaluating these approaches. Lastly, we identify key challenges for clinical implementation of the best multicentric harmonization strategy identified in our experiments for enabling quantitative neuroradiological assessment of Alzheimer’s disease (QNAD).

## 2 Materials and Methods

### 2.1 Participants and scanner characteristics

T1w MRI data of healthy controls (CN), participants with subjective cognitive decline (SCD), mild cognitive impairment (MCI), and Alzheimer’s disease (AD) from 11 data cohorts were included in our analysis. The cohorts considered for this study were: Amsterdam Dementia Cohort (ADC)^16^, Alzheimer’s Disease Neuroimaging Initiative (ADNI)^17^, Australian Imaging, Biomarker & Lifestyle Flagship Study of Ageing (AIBL)^18^, Alzheimer’s Repository Without Borders (ARWiBo)^19^, European DTI Study on Dementia (EDSD)^20^, Hungarian Longitudinal Study of Healthy Brain Aging (HuBA)^21^, Italian Alzheimer’s Disease Neuroimaging Initiative (I-ADNI)^22^, National Alzheimer’s Coordination Center (NACC) ^23^, Open Access Series of Imaging Studies (OASIS, versions 1&2)^24^, European Alzheimer’s Disease Neuroimaging Initiative (also known as PharmaCOG)^25^, and UK Bio-bank (UKBB)^26^. Detailed information about each cohort is summarized in Supplementary Table 1.

Minimum inclusion criteria included the availability of a T1w MRI scan along with age, sex, scanner information, and a clinical diagnosis of either CN, SCD, MCI or AD. All datasets were organized according to the BIDS standard^27^ to ensure inter-operability and data anonymization. An overview of the scanners used in this study is shown in Table 1.

**Table 1:**
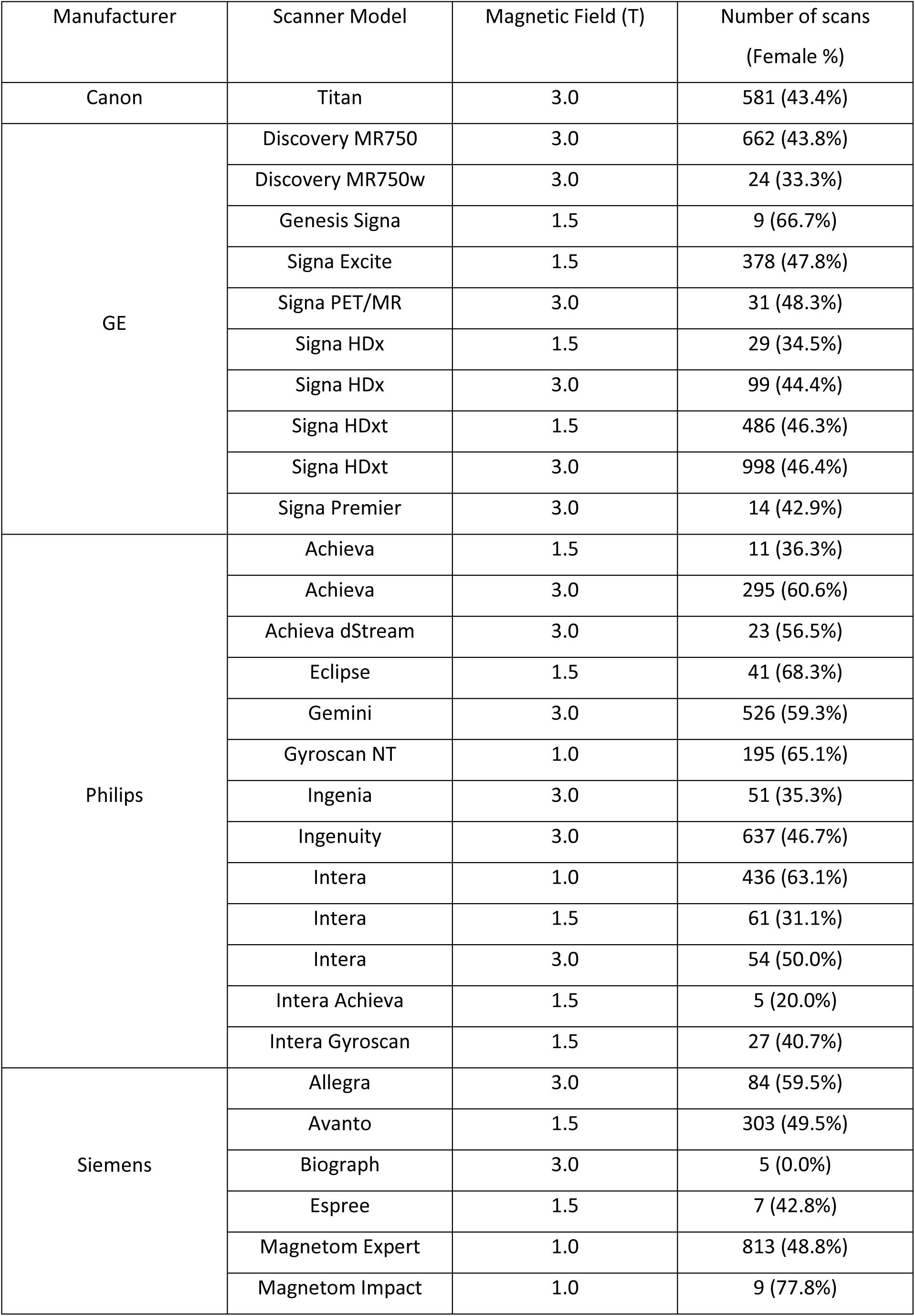
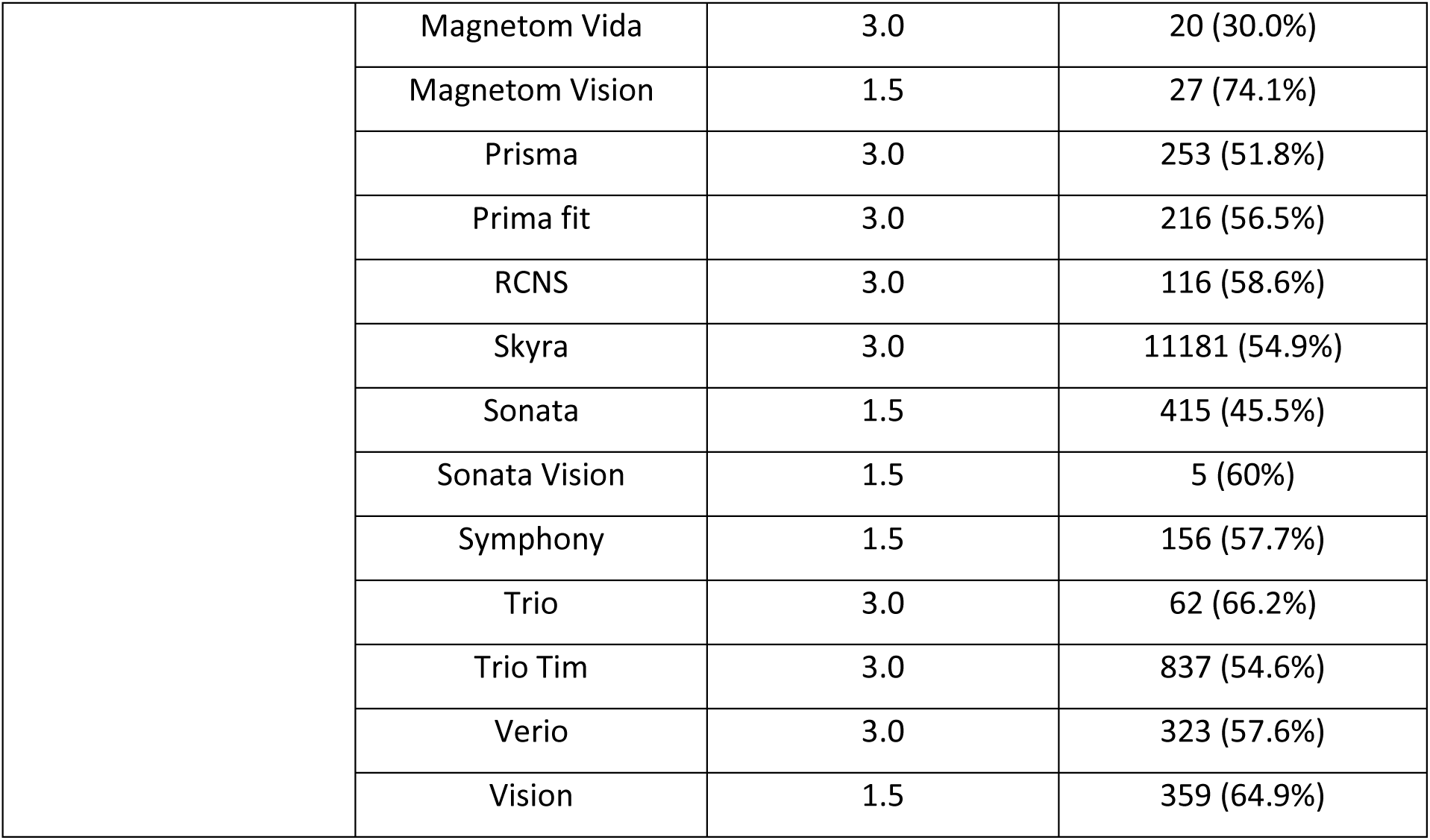
Scanners considered in this study and their characteristics.

### 2.2 Image processing

Cortical reconstruction and volumetric segmentation were performed with the cross-sectional pipeline of FreeSurfer v7.1.1^28^ in order to extract volumes of 68 cortical regions in the Desikan-Killiany atlas, 14 subcortical brain regions, as well as total CSF volume, total gray matter volume and total brain volume with and without ventricles. All features derived from FreeSurfer are listed in Supplementary Figure 1. IQM were estimated using MRIQC v0.16.1^29^. Automatic quality control of the FreeSurfer segmentations was performed using the Euler number, where outliers defined as 1.5×IQR (inter-quartile range) below the first quartile and 1.5×IQR above the third quartile^30^ per scanner were excluded from our experiments.

In order to ensure reproducibility of our results across different computing environments^31^, Docker containers for both FreeSurfer (https://github.com/E-DADS/freesurfer) and MRIQC (https://github.com/E-DADS/mriqc) have been made available online.

### 2.3 Multi-class Neuroharmony model

The volumetric data from all individuals in the training set were harmonized using ComBat harmonization^10^ with empirical Bayes optimization, for removing scanner related batch effects while preserving the effects of age, sex, and cognitive status. The cognitive status was dichotomized based on the clinical diagnosis as either CU (CN and SCD) or CI (MCI and AD). Subsequently, a random forest regressor was trained with MRIQC-derived IQM to predict the corrections needed to harmonize the volumes as predicted by ComBat. To ensure the regressor learns to predict the corrections needed to harmonize accurately in the presence of AD pathology, we used synthetic minority oversampling technique (SMOTE) for data augmentation^32^ before training the random forest regressor. This ensured that IQM values with and without neurodegeneration were equally distributed by removing data imbalance between CI and CU individuals. The use of dichotomized cognitive status instead of clinical diagnosis ensured that in the test phase, a full clinical diagnosis is not required as an input to predict the harmonized volumes. The hyperparameters for the random forest regressor were chosen to be the same as the ones used in the original Neuroharmony paper.^15^

### 2.4 Model comparisons

The performance of the proposed multi-class extension to Neuroharmony was compared with two other machine-learning based harmonization strategies that are generalizable to external datasets:

#### 2.4.1 Normative model

In the training phase, volumetric data from only the CU individuals were harmonized using ComBat harmonization using the aforementioned strategy, while preserving the effects of age and sex. Subsequently, a random forest regressor was trained to predict the corrections needed to harmonize the volumes as predicted by ComBat using MRIQC-derived IQM.

#### 2.4.2 Inclusive model

The training strategy remained the same as for the normative model, but volumetric data of both CU and CI individuals were used for training.

### 2.5 Measures for model evaluations

We use two evaluation metrics to assess the ability of the harmonization strategies to remove scanner-related variations in brain-volumetric data between each pair of scanners (biomarker concordance), while retaining the ability to delineate diagnostic groups (preserving disease-related signal).

Firstly, to assess if the volumetric data are harmonized, we compared the distributions of each volumetric measure for each pair of scanners. This was done independently within each diagnostic group, and after correcting for the confounding effects of age and sex by regressing out their effects estimated in CU individuals. The Kolmogorov-Smirnov (KS) test was used for comparing these distributions with the null hypothesis that the distributions between any two pair of scanners were the same. For statistical validity we excluded scanners with fewer than 10 participants of the same diagnostic group from this evaluation. A measure for evaluating harmonization was defined as the percentage of such comparisons across brain regions for each scanner pair, where distributions were *not* statistically different from each other (p ≥ 0.05) after correcting for multiple testing via false discovery rate (FDR). To provide a reference measure for biomarker concordance, we also computed this measure in the non-harmonized data.

Secondly, to assess if disease-related signal was preserved in the volumetric measures, we used area under the receiver operating characteristic (ROC) curve (AUC) as an auxiliary evaluation measure. The ROC curve for distinguishing CN participants from AD patients was computed independently for each volumetric measure. A reference measure for AUC was also computed for the non-harmonized dataset.

### 2.6 Cross-validation experiments

We performed two experiments in a cross-validation framework. Experiment 1 assessed concordance of the three harmonization strategies, by performing cross-validation at the scanner-level. Experiment 2 performed cross-validation at the participant level, using the best-performing scanner-level harmonization models.

Experiment 1: To investigate the generalizability of the model to unseen scanners (not included in the training set), we performed 5-fold cross-validation across the 43 available scanners, where in each fold 80% of the scanners were used for training the models, and the remaining 20% of the scanners were used for evaluation. In this experiment, to evaluate the bias introduced by using single-scanner data from the large UKBB cohort, we repeated this experiment for increasing portions of UKBB participants such that when the UKBB data is included in the training data the proportions included were: 10%; 33%; 67%; 100%. However, when UKBB cohort data is used in the test set, we always used 100% of the cohort. The non-parametric McNemar Chi-square test was used to compare accuracies across harmonization strategies.

Experiment 2: In this experimental setup, the test set consisted of participants from the scanners which were also present in the training set. We selected the two best performing models from Experiment 1 and performed a stratified 5-fold cross-validation across participants, stratified based on the dichotomized cognitive status. For this, the proportion of the UKBB participants included was also decided based on Experiment 1. To provide a reference measure, we compared the accuracies obtained with the corresponding accuracies obtained in Experiment 1.

## 3 Results

### 3.1 Participants

Table 2 shows descriptive statistics for the combined study sample used in our experiments, which consisted of QC-passed volumetric data from 20,864 participants (53.3% female) from 43 scanners across 11 cohorts. Figure 1 shows age distributions by scanner and cognitive group.

**Figure 1:**
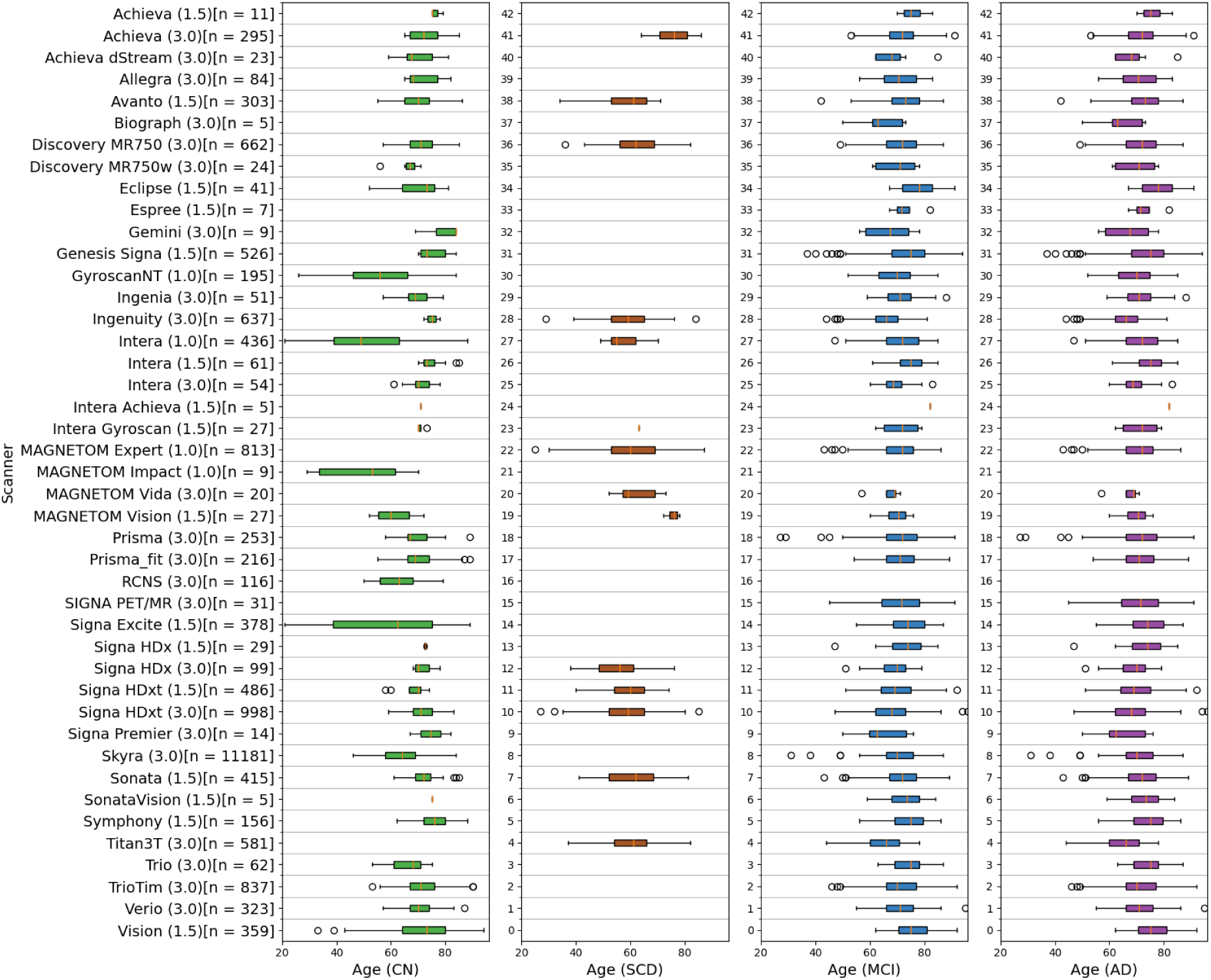
Diagnosis-wise distributions of age for each scanner in the training cohort. Abbreviations: CN: cognitively normal; SCD: subjective cognitive decline; MCI: mild cognitive impairment; AD: Alzheimer’s Disease.

**Table 2:** Participant Demographics. † Values indicated in the column are calculated after removing the outliers, as described in section 2.2. Abbreviations: CN: cognitively normal; SCD: subjective cognitive decline; MCI: mild cognitive impairment; AD: Alzheimer’s Disease.

|  | Participants<br>(processed/<br>considered<br>after removing<br>outliers) | Age [years]<sup>†</sup> | Sex (F/M)<sup>†</sup> | Diagnosis<br>(CN / SCD / MCI / AD)<sup>†</sup> | Unique<br>scanners<sup>†</sup> |
| --- | --- | --- | --- | --- | --- |
| ADC | 4,086 / 3,722 | 63.9 ± 9.2 | 1,717 / 2,005 | 0 / 1,355 / 805 / 1562 | 12 |
| ADNI | 2,044 / 1,830 | 72.2 ± 7.06 | 889 / 941 | 687 / 0 / 851 / 292 | 27 |
| AIBL | 557 / 524 | 72.7 ± 6.5 | 299 / 225 | 388 / 0 / 83 / 53 | 3 |
| ARWiBo | 913 / 831 | 56.3 ± 16.2 | 529 / 302 | 603 / 16 / 116 / 96 | 7 |
| EDSD | 416 / 384 | 70.4 ± 7.3 | 197 / 187 | 143 / 0 / 119 / 122 | 8 |
| HuBA | 121 / 116 | 62.4 ± 6.9 | 68 / 48 | 116 / 0 / 0 / 0 | 1 |
| I-ADNI | 179 / 172 | 72.2 ± 8.0 | 106 / 66 | 2 / 5 / 35 / 130 | 4 |
| NACC | 1,861 / 1,731 | 71.9 ± 9.8 | 910 / 821 | 0 / 0 / 949 / 782 | 22 |
| OASIS | 373 / 359 | 73.2 ± 10.7 | 233 / 126 | 211 / 0 / 111 / 37 | 1 |
| PharmaCog | 141 / 137 | 69.0 ± 7.3 | 80 / 57 | 0 / 0 / 137 / 0 | 7 |
| UKBB | 12,259 /<br>11,058 | 63.5 ± 7.6 | 6,083 / 4,975 | 11,058 / 0 / 0 / 0 | 1 |
| Total | 22,950 /<br>20,864 | 65.3 ± 9.4 | 11,111 /<br>9,753 | 13,208 / 1,376 / 3,206 /<br>3,074 | 43 |

### 3.2 Model evaluation

Figure 2 shows the first result of Experiment 1: biomarker concordance under cross-validation, independently for each diagnostic group and with increasing proportions of the UKBB dataset. Reference concordance for non-harmonized dataset are also shown for each diagnostic group for comparison. As expected, concordance for each harmonization strategy were significantly higher than the non-harmonized data for all the diagnostic groups (*p* < 10^−6^). The use of the inclusive and multi-class models significantly improved the concordance with respect to the normative model for the diagnostic categories of MCI and AD (*p* < 10^−6^). For diagnostic groups of CN and SCD, the concordance of the multi-class model was significantly higher than the normative model when the proportion of UKBB subjects included was 100% (*p*_*CN*_ = 0.01, *p*_*SCD*_ = 0.02). The inclusive model’s concordance for CN and SCD subjects was statistically similar to that of the normative model (*p* > 0.05).

**Figure 2:**
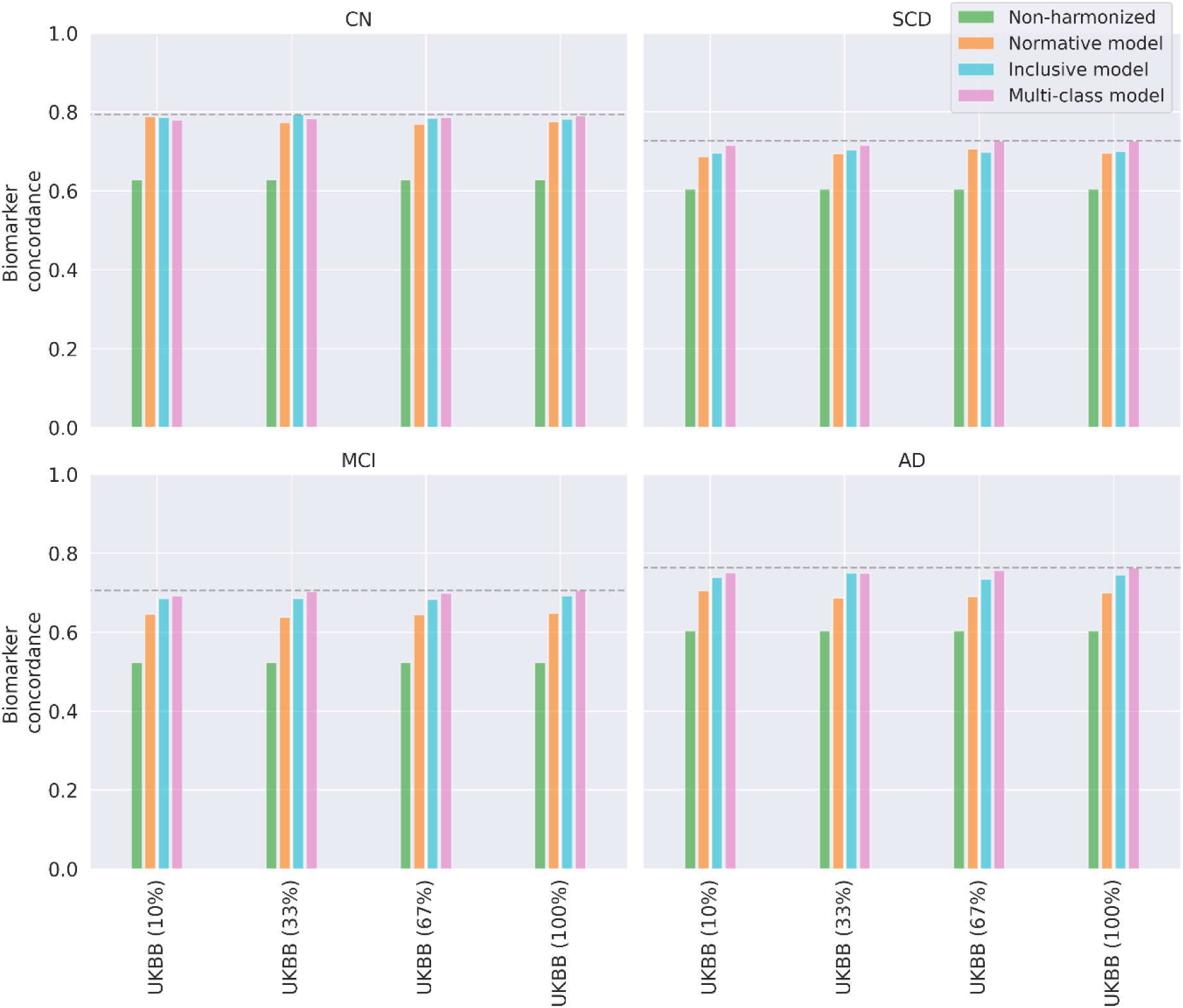
Experiment 1: Biomarker concordance for brain volumes on unseen scanners using different harmonization strategies. Concordance for non-harmonized data has also been shown here as a reference measure for comparison. Abbreviations: CN: cognitively normal; SCD: subjective cognitive decline; MCI: mild cognitive impairment; AD: Alzheimer’s Disease.

Figure 3 shows the second result of Experiment 1: CN vs AD AUC computed independently for each brain regional volume in the test set. Removing scanner-related differences decreased AUC for all harmonization approaches, potentially due to the imbalance in the number of CN and AD participants in the different scanners. The AUCs of the normative model and multi-class model are slightly lower than non-harmonized volumes (*ΔAUC* = −0.01, paired t-test *p* = 10^−5^). However, the multi-class model significantly outperformed the inclusive model (*ΔAUC* = −0.09, paired t-test *p* < 10^−10^), indicating relative loss of disease related signal when using the inclusive model harmonization strategy.

**Figure 3:**
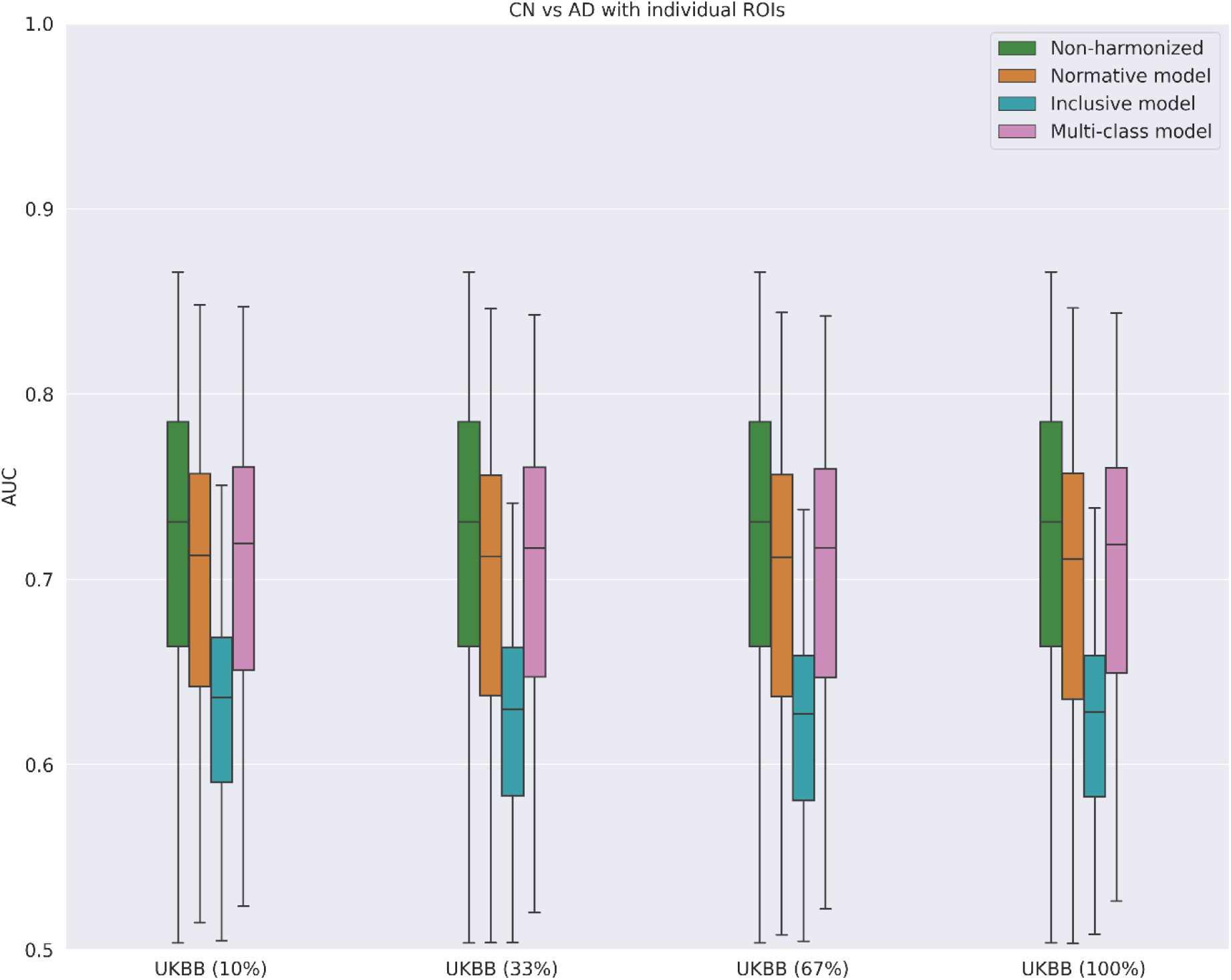
Experiment 1: Boxplots of AUCs for distinguishing CN participants from AD patients in the test set based on the 86 brain ROIs considered before and after harmonization. Abbreviations: CN: cognitively normal; AD: Alzheimer’s Disease.

### 3.3 Harmonization in seen vs unseen MRI scanners

Figure 4 shows the results of Experiment 2: biomarker concordance as a function of scanners seen vs unseen scanners during model training, for the normative model and the multi-class model. Supplementary Figure 1 shows the same for each brain volume individually. Biomarker concordance of the multi-class model was significantly higher than the normative model for unseen scanners, based on McNemar test for all diagnostic categories (*p*_*CN*_ = 0.01, *p*_*SCD*_ = 0.02, *p_MCI_* < 10^−6^, *p_AD_* < 10^−6^). For seen scanners, the multi-class model harmonization strategy significantly outperformed the normative model for the diagnostic groups of CN, MCI, and AD (*p*_*CN*_ < 10^−6^, *p_MCI_* < 10^−6^, *p_AD_* < 10^−6^), but significantly underperformed for SCD (*p*_*SCD*_ = 0.02). Harmonization accuracy using the multi-class model in a seen scanner (accuracy > 97%) was better for all diagnostic groups than in unseen scanners (accuracy < 79%).

**Figure 4:**
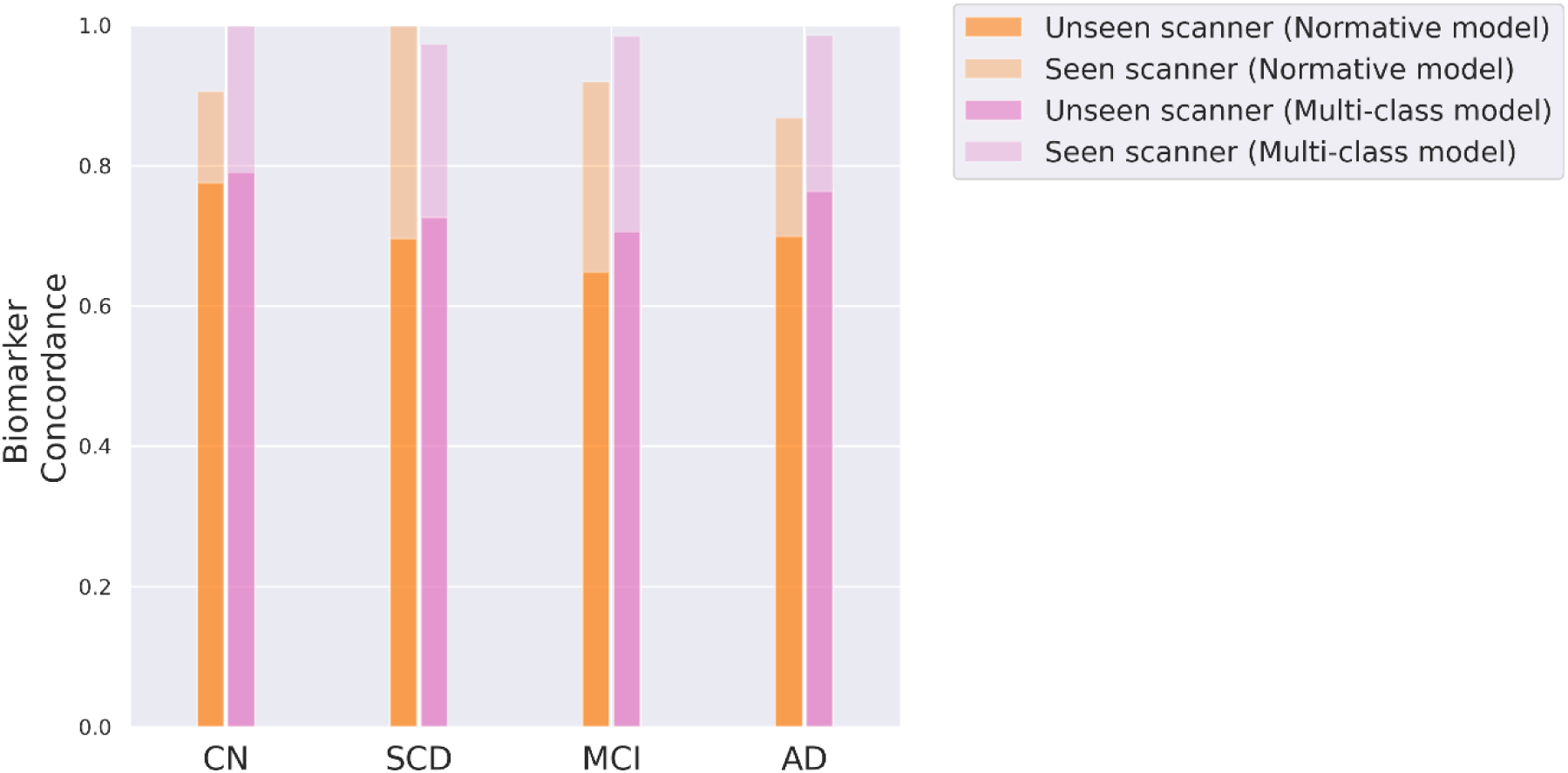
Experiment 2: Accuracy of harmonization for harmonizing brain volumes on seen versus unseen scanners using normative model and multi-class model. Abbreviations: CN: cognitively normal; SCD: subjective cognitive decline; MCI: mild cognitive impairment; AD: Alzheimer’s Disease.

## 4 Discussion

We introduced a novel extension to the Neuroharmony harmonization model to train a generalizable machine learning model for harmonizing multicentric brain-volumetric data for quantitative neuro-radiological assessment of Alzheimer’s disease. The data for these evaluation experiments were derived from T1w MRIs acquired with 43 different scanners from 20,864 participants spanning 11 cohorts. The newly introduced multi-class model would be helpful in harmonizing volumetric data while using automated methods in clinics and research where there could be data from new scanners not included in training. The trained model has been made openly available at https://www.neugrid2.eu/index.php/edads_harmonization/ while the code to train the model has been made available in: https://github.com/e-dads/Multiclass-Neuroharmony/.

Our experiments showed that the multi-class model, that accounts for the interaction between disease pathology and image quality metrics to remove scanner-related effects, significantly improved harmonization accuracy for patients in unseen scanners, as compared to normative modelling. For seen scanners, it improved the harmonization accuracy for all diagnostic groups except SCD, potentially due to the low sample size of the SCD group (n=1,376). Additionally, we showed that the multi-class model of Neuroharmony preserves disease-related signature during harmonization.

Harmonization of biomarker data from unseen scanners remains a challenge: biomarker concordance for both normative and multi-class models in unseen scanners was lower than obtained for seen scanners. While this leaves scope for further methodological improvements to harmonization strategies for unseen scanners, it would also be useful to investigate if the achieved harmonization performance is sufficient for the generalizability of machine learning approaches such as classification, subtyping^33^, and brain aging.

The different number of participants used to train the respective models could potentially bias the results against the model which uses a smaller dataset for training (normative model). However, we think this setting is a realistic and fair comparison, because normative modelling always discards data from CI individuals. Through our modifications to the Neuroharmony model, we provided a way to include both CI and CU individuals in the training data, which our experiments show improves harmonization in both seen and unseen scanners while preserving disease-related signature.

The harmonization performance obtained with the normative model in our experiments was lower than reported in the original Neuroharmony paper^15^. We think this is due to removal of sex and age variability in the original Neuroharmony method. We preserved these effects, retaining this biological variability, which we would argue is important for both research studies and future clinical implementation.

Some limitations of the original Neuroharmony model^15^ also apply to this work as well. The harmonization performance for an individual in the test-set depends on the contrast-to-noise ratio in the T1w MRI and the pipeline cannot guarantee effective harmonization if the ratio is outside the range seen in our training data, and might lead to incorrect harmonization. Secondly, the harmonization performance based on biomarker concordance across scanner-pairs is a surrogate measure to measure consistency in the absence of a ground-truth. To overcome this limitation, it would be useful to measure the harmonization performance for participants scanned with multiple scanners, in the future.

An important limitation of this study, as with most research studies in this field, is that the imaging data used predominantly came from the developed Western countries of the EU, US, UK, and Australia. A more generalizable and inclusive machine learning model for harmonization would require data from nations in South-America, Asia, and Africa. This would include low field-strength scanners that are predominantly used in these regions, as well as more diverse biological variation in the training data. Large global consortia such as the UNITED consortium^34^ could potentially help in getting access to such diverse neuroimaging data. Further developing Neuroharmony for distributed or federated learning for harmonizing imaging data can also facilitate data inclusion from under-represented countries.

Challenges in the clinical implementation of the harmonization strategy: while the multi-class model outperformed the normative model in terms of harmonization accuracy, the implementation of the model in memory clinics might require additional work to include cognitive status of a patient during regular radiological workup. Machine learning models could potentially be used to overcome this limitation as it has been shown in recent studies that classifying CI from CN/SCD can be done with high accuracy using MRI^35^. To avoid a circular dependency between the two tasks, we think that developing multi-task machine learning models to jointly harmonize and predict cognitive status is an important avenue of future work.

While the current work was focused on the AD spectrum, we expect that our new method will be valuable for impaired cognition in general (e.g.: vascular dementia, frontotemporal dementia, dementia with Lewy bodies). Future work validating harmonization approaches for patients with other types of dementia is crucial for eventual clinical implementation.

In summary, we have generalized the Neuroharmony model to harmonize imaging biomarker data from multisite studies while retaining disease signal that could otherwise be removed by the harmonization procedure. Demonstrating on brain MRI biomarker data from the Alzheimer’s disease spectrum, our new method outperforms others on both seen and unseen scanners, making it more suitable for clinical applications related to cognitive decline, such as memory clinics and clinical trials of new interventions for neurodegenerative diseases.

## Data Availability

ADC data can be made available to academic researchers upon reasonable request;
ADNI and AIBL data are stored at the Laboratory of Neuroimaging at the University of Southern California and are available to the general scientific community for download: http://adni.loni.usc.edu;
ArWiBO, EDSD, I-ADNI, OASIS and PharmaCog data are available for all researchers on the NeuGRID2 platform: https://www.neugrid2.eu/ (https://doi.org/10.17616/R31NJN1E);
HuBA data can be made available upon reasonable request;
NACC data is available through the National Alzheimer’s Coordinating Center platform: https://naccdata.org/;
UKBB data is available at the UK Biobank platform: https://www.ukbiobank.ac.uk/;

http://adni.loni.usc.edu

https://www.neugrid2.eu

https://naccdata.org

https://www.ukbiobank.ac.uk

## Acknowledgments

The authors would like to thank the developers of the original Neuroharmony algorithm for making the code available, and all the research participants and their families for donating their data for scientific research.

Data collection and sharing for ADNI was funded by the Alzheimer’s Disease Neuroimaging Initiative (ADNI) (National Institutes of Health Grant <u>U01 AG024904</u>) and DOD ADNI (Department of Defense award number <u>W81XWH-12-2-0012</u>). ADNI is funded by the National Institute on Aging, the National Institute of Biomedical Imaging and Bioengineering, and through generous contributions from the following: AbbVie, Alzheimer’s Association; Alzheimer’s Drug Discovery Foundation; Araclon Biotech; BioClinica, Inc.; Biogen; Bristol-Myers Squibb Company; CereSpir, Inc.; Cogstate; Eisai Inc.; Elan Pharmaceuticals, Inc.; Eli Lilly and Company; EuroImmun; F. Hoffmann-La Roche Ltd and its affiliated company Genentech, Inc.; Fujirebio; GE Healthcare; IXICO Ltd.; Janssen Alzheimer Immunotherapy Research & Development, LLC.; Johnson & Johnson Pharmaceutical Research & Development LLC.; Lumosity; Lundbeck; Merck & Co., Inc.; Meso Scale Diagnostics, LLC.; NeuroRx Research; Neurotrack Technologies; Novartis Pharmaceuticals Corporation; Pfizer Inc.; Piramal Imaging; Servier; Takeda Pharmaceutical Company; and Transition Therapeutics. The Canadian Institutes of Health Research is providing funds to support ADNI clinical sites in Canada. Private sector contributions are facilitated by the Foundation for the National Institutes of Health (www.fnih.org). The grantee organization is the Northern California Institute for Research and Education, and the study is coordinated by the Alzheimer’s Therapeutic Research Institute at the University of Southern California. ADNI data are disseminated by the Laboratory for Neuro Imaging at the University of Southern California.

The AIBL study (https://aibl.org.au) is a consortium between Austin Health, CSIRO, Edith Cowan University, the Florey Institute (The University of Melbourne), and the National Ageing Research Institute. The study has received partial financial support from the Alzheimer’s Association (US), the Alzheimer’s Drug Discovery Foundation, an Anonymous foundation, the Science and Industry Endowment Fund, the Dementia Collaborative Research Centres, the Victorian Government’s Operational Infrastructure Support program, the Australian Alzheimer’s Research Foundation, the National Health and Medical Research Council (NHMRC), and The Yulgilbar Foundation. Numerous commercial interactions have supported data collection and analyses. In-kind support has also been provided by Sir Charles Gairdner Hospital, Cogstate Ltd, Hollywood Private Hospital, The University of Melbourne, and St Vincent’s Hospital. The AIBL team wishes to thank all clinicians who referred patients with AD and/or MCI to the study. We also thank all those who took part as subjects in the study for their commitment and dedication to helping advance research into the early detection and causation of AD. We thank all the investigators within the AIBL who contributed to the design and implementation of the resource and/or provided data but did not actively participate in the development, analysis, interpretation or writing of this current study.

The NACC database is funded by NIA/NIH Grant U24 AG072122. NACC data are contributed by the NIA-funded ADRCs: P30 AG062429 (PI James Brewer, MD, PhD), P30 AG066468 (PI Oscar Lopez, MD), P30 AG062421 (PI Bradley Hyman, MD, PhD), P30 AG066509 (PI Thomas Grabowski, MD), P30 AG066514 (PI Mary Sano, PhD), P30 AG066530 (PI Helena Chui, MD), P30 AG066507 (PI Marilyn Albert, PhD), P30 AG066444 (PI John Morris, MD), P30 AG066518 (PI Jeffrey Kaye, MD), P30 AG066512 (PI Thomas Wisniewski, MD), P30 AG066462 (PI Scott Small, MD), P30 AG072979 (PI David Wolk, MD), P30 AG072972 (PI Charles DeCarli, MD), P30 AG072976 (PI Andrew Saykin, PsyD), P30 AG072975 (PI David Bennett, MD), P30 AG072978 (PI Neil Kowall, MD), P30 AG072977 (PI Robert Vassar, PhD), P30 AG066519 (PI Frank LaFerla, PhD), P30 AG062677 (PI Ronald Petersen, MD, PhD), P30 AG079280 (PI Eric Reiman, MD), P30 AG062422 (PI Gil Rabinovici, MD), P30 AG066511 (PI Allan Levey, MD, PhD), P30 AG072946 (PI Linda Van Eldik, PhD), P30 AG062715 (PI Sanjay Asthana, MD, FRCP), P30 AG072973 (PI Russell Swerdlow, MD), P30 AG066506 (PI Todd Golde, MD, PhD), P30 AG066508 (PI Stephen Strittmatter, MD, PhD), P30 AG066515 (PI Victor Henderson, MD, MS), P30 AG072947 (PI Suzanne Craft, PhD), P30 AG072931 (PI Henry Paulson, MD, PhD), P30 AG066546 (PI Sudha Seshadri, MD), P20 AG068024 (PI Erik Roberson, MD, PhD), P20 AG068053 (PI Justin Miller, PhD), P20 AG068077 (PI Gary Rosenberg, MD), P20 AG068082 (PI Angela Jefferson, PhD), P30 AG072958 (PI Heather Whitson, MD), P30 AG072959 (PI James Leverenz, MD).

## Data and code availability

ADC data can be made available to academic researchers upon reasonable request;

ADNI and AIBL data are stored at the Laboratory of Neuroimaging at the University of Southern California and are available to the general scientific community for download: http://adni.loni.usc.edu;

ArWiBO, EDSD, I-ADNI, OASIS and PharmaCog data are available for all researchers on the NeuGRID2 platform: https://www.neugrid2.eu/ (https://doi.org/10.17616/R31NJN1E);

HuBA data can be made available upon reasonable request;

NACC data is available through the National Alzheimer’s Coordinating Center platform: https://naccdata.org/;

UKBB data is available at the UK Biobank platform: https://www.ukbiobank.ac.uk/;

Docker containers for FreeSurfer and MriQC are available on the E-DADS GitHub: https://github.com/E-DADS/freesurfer, https://github.com/E-DADS/mriqc;

Multi-class Neuroharmony harmonization algorithm is available on GitHub: https://github.com/88vikram/Multiclass-Neuroharmony;

Harmonization model files are available for all researchers on the NeuGRID2 platform: https://www.neugrid2.eu/index.php/edads_harmonization.

