## Supplemental material for "A machine-learning model to harmonize brain volumetric data for quantitative neuro-radiological assessment of Alzheimer’s disease"

### Supplementary material

| Cohort | Full name | Description | Categories |
|---|---|---|---|
| ADC | Amsterdam Dementia Cohort | The ADC includes all patients who come to the Alzheimer Center in Amsterdam (since 2004) for diagnostic work-up and consent to give all their data collected for research. The aim is to facilitate research into new and existing biomarkers in the broadest sense, to establish diagnostic, prognostic values, and further insight into the pathogenesis of neurodegenerative dementias. The data consist of baseline and annual follow-up assessments. Clinical, neuropsychological, imaging, and biological markers are collected. Since it is conception, it has grown into one of the largest clinical data sets in the dementia field. | SCD<br>MCI<br>AD |
| ADNI | Alzheimer’s Disease Neuroimaging Initiative | The Alzheimer’s Disease Neuroimaging Initiative is a longitudinal multicentre study designed to develop clinical, imaging, genetic, and biochemical biomarkers for the early detection and tracking of Alzheimer’s disease (AD). ADNI was originally launched in 2003 as a public-private partnership; its primary goal has been to test whether magnetic resonance imaging (MRI), biological markers, clinical and neuropsychological assessments can be combined to measure the progression of MCI and Alzheimer's disease. The initial five-year study (ADNI-1) was extended by two years in 2009 by a Grand Opportunities grant (ADNI-GO), and in 2011 by further competitive renewal of the ADNI-1 grant (ADNI-2). Through its 3 phases, it has targeted participants with AD, different stages of MCI, and CN. | CN<br>MCI<br>AD |
| AIBL | | The Australian Imaging, Biomarker & Lifestyle Flagship Study of Ageing is a study to discover which biomarkers, cognitive characteristics, and health and lifestyle factors | CN<br>MCI<br>AD |

| | | | |
|---|---|---|---|
| | | determine subsequent development of symptomatic Alzheimer’s Disease (AD). | |
| ARWiBo | Alzheimer Disease Repository Without Borders | ARWiBo is a cross-sectional data set including data from more than 2,500 patients enrolled in Brescia (Italy) and nearby areas. The data set contains socio-demographic, clinical, genotype, bio-specimen information, MRI T1-weighted images. | CN<br>SCD<br>MCI<br>AD |
| EDSD | European DTI Study on Dementia | EDSD is a framework of nine European centres: Amsterdam (Netherlands), Brescia (Italy), Dublin (Ireland), Frankfurt (Germany), Freiburg (Germany), Milano (Italy), Mainz (Germany), Munich (Germany), and Rostock (Germany). It is a cross-sectional multi-centre study characterized by 474 volumetric MRI T1-weighted scans with socio-demographic, clinical, genetic, and biological variables. | CN<br>MCI<br>AD |
| HuBA | Hungarian Longitudinal Study of Healthy Brain Aging | Hungarian adaptation of the Australian Imaging, Biomarkers and Lifestyle (AIBL) study of aging study protocol. It is a collection of demographic, lifestyle, mental and physical health, medication and medical history related information as well as series of magnetic resonance imaging (MRI) data. Participants were also offered to participate in collection of blood samples as well as in a sleep study aimed at assessing the general sleep quality based on multi-day acquisition of subjective sleep questionnaires and whole-night electroencephalographic (EEG) data. | CN |
| I-ADNI | Italian ADNI | I-ADNI is a cross sectional study and consists of 262 patients with subjective memory impairment, mild cognitive impairment, Alzheimer's dementia, and frontotemporal dementia enrolled in 7 Italian centers. Few cognitively healthy elderly controls were also included. MRI site qualification and MP-RAGE quality assessment was applied following the NA-ADNI procedures. | CN<br>SCD<br>MCI<br>AD |

| | | | |
|---|---|---|---|
| NACC | National Alzheimer’s Coordinating Center | NACC is home to one of the largest, oldest, and most powerful Alzheimer’s datasets, built in collaboration with more than 42 Alzheimer’s Disease Research Centers (ADRCs) throughout the US over the past 20+ years. | MCI<br>AD |
| OASIS | Open Access Series of Imaging Studies | OASIS consists of (I) a cross-sectional collection of 416 subjects. 100 of the included subjects, over the age of 60, have been clinically diagnosed with very mild to moderate Alzheimer's disease (AD). (II) A longitudinal collection of 150 subjects aged from 60 to 96 years. Each subject was scanned on two or more visits, separated by at least one year for a total of 373 imaging sessions. In addition, the data set contains socio-demographic, clinical, genotype information. | CN<br>MCI<br>AD |
| PharmaCog (E-ADNI) | Prediction of cognitive properties of new drug candidates for neurodegenerative diseases in early clinical development | PharmaCog is an industry-academic European project (IMI) aimed at identifying biomarkers sensitive to symptomatic and disease modifying effects of drugs for Alzheimer's disease. Several clinical sites participated in this study across Italy (Brescia, Verona, Milan, Perugia, and Genoa), Spain (Barcelona), France (Marseille, Lille, and Toulouse), Germany (Leipzig and Essen), Greece (Thessaloniki) and Netherland (Amsterdam). 151 MCI patients have been studied longitudinally collecting multimodal image scans, clinical variables, and bio-specimens. | MCI |
| UKBB | | UK Biobank is a large-scale biomedical database and research resource, containing in-depth genetic and health information from half a million UK participants. The database is regularly augmented with additional data and is globally accessible to approved researchers undertaking vital research into the most common and life-threatening diseases. It is a major contributor to the advancement of modern medicine and treatment and | CN |

| | | |
|---|---|---|
| | | has enabled several scientific discoveries that improve human health. |

**Supplementary Table 1:** Cohort details. Acronyms: CN: cognitively normal; SCD: subjective cognitive decline; MCI: mild cognitive impairment; AD: Alzheimer's Disease.

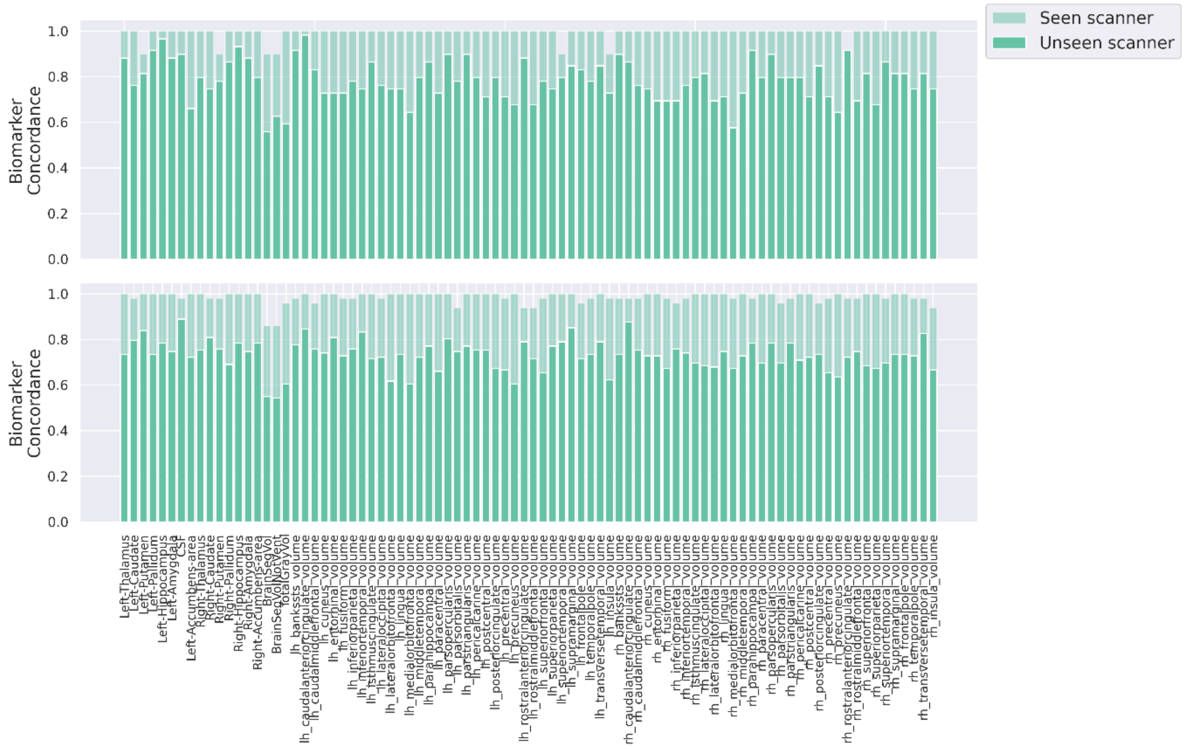

**Supplementary Figure 1:** Experiment 2: Harmonization accuracy for seen scanners and unseen scanners for each volumetric feature for cognitively unimpaired participants (top) and patients with cognitive impairment (bottom).
